# Weighted analysis of symptom profile by vaccination status in positive SARS-CoV-2 cases: observational study in São Gonçalo, Brazil

**DOI:** 10.1101/2024.12.03.24318348

**Authors:** Raphael Rangel das Chagas, Hércules Rezende Freitas, Sergian Vianna Cardozo

## Abstract

This study investigates symptom associations in SARS-CoV-2 positive individuals based on vaccination status. Unvaccinated individuals exhibited significantly higher odds of experiencing severe symptoms, including fever > 38.5□°C (_log_OR = 3.64, 95% CI: 1.52–5.77, p = 0.0008), rhinitis (_log_OR = 2.94, 95% CI: 0.82–5.05, p = 0.0065), headache (_log_OR = 2.17, 95% CI: 0.06–4.28, p = 0.0436), and myalgia (_log_OR = 3.25, 95% CI: 1.17–5.34, p = 0.0023). Conversely, unvaccinated individuals were less likely to report cough (_log_OR = -2.44, 95% CI: -4.53 to -0.34, p = 0.0226), potentially reflecting behavioral factors related to vaccine hesitancy. Among vaccinated participants, symptom profiles varied by vaccine type. Both Oxford-AstraZeneca and Pfizer-BNT162b2 vaccines were associated with increased odds of loss of smell (_log_OR = 0.81 and 1.47, respectively) and loss of taste (_log_OR = 0.75 for Oxford-AstraZeneca). Additionally, the Oxford-AstraZeneca vaccine was linked to dyspnea (_log_OR = 0.85, 95% CI: 0.02–1.69, p = 0.0449). These findings suggest that vaccination reduces the likelihood of severe systemic and respiratory symptoms while influencing specific symptom manifestations in breakthrough cases. Ongoing research is essential to understand vaccine-specific immune responses and their impact on clinical outcomes.

## Introduction

The rapid development and deployment of vaccines during the SARS-CoV-2 pandemic played a critical role in mitigating the COVID-19 crisis. Vaccines such as BNT162b2 (Pfizer-BioNTech) demonstrated high efficacy in preventing symptomatic disease, severe outcomes, and hospitalizations, particularly during initial rollouts (Polack et al. 2020). This success was enabled by global collaborations, accelerated trials, and regulatory adaptations that maintained safety while reducing traditional development timelines. Vaccines also reduced transmission rates and viral loads, indirectly protecting vulnerable individuals (Baden et al. 2021).

High vaccine coverage curbed the spread of variants of concern, though disparities in access highlighted inequities in low-income regions (Lurie et al. 2020). Public health communication was pivotal in addressing vaccine hesitancy, a key barrier to herd immunity (MacDonald 2015). The pandemic underscored the importance of investing in vaccine research infrastructure to enable rapid responses to emerging threats, emphasizing preparedness for future public health emergencies.

The goal of this short communication is to contribute to the growing body of knowledge about vaccine safety, a foundation that will be crucial for managing future public health crises such as the COVID-19 pandemic.

## Materials and methods

### Experimental design

This observational, cross-sectional, and analytical study is based on our previous work (Chagas et al. 2024, under review). Briefly, patients who sought care at COVID-19 screening centers in São Gonçalo, Rio de Janeiro, Brazil, between July 28 and December 2, 2021, were invited to enroll in the study. The sample data were weighted to align with São Gonçalo’s demographics, which in 2022 included a population of 896,744, a density of 3,613.57 inhabitants per km^2^, and a total area of 248.16 km^2^. Socioeconomic and health indicators for the municipality include a 96.7% school enrollment rate among children aged 6–14 (2010), a Municipal Human Development Index (MHDI) of 0.739, a GDP per capita of R$18,504.81 (2021), and an infant mortality rate of 12.33 per 1,000 live births (2020) (IBGE 2024).

### Inclusion criteria and informed consent

Patients in this study exhibited symptoms suggestive of SARS-CoV-2 infection and underwent COVID-19 testing using the RT-LAMP technique at screening centers in São Gonçalo, Rio de Janeiro. Adhering to the Declaration of Helsinki and relevant Brazilian legislation, the study prioritized ethical integrity, ensuring confidentiality by anonymizing all data prior to analysis. Approved by the Research Ethics Committee of the University of Grande Rio (CEP/UNIGRANRIO) under CAAE number 32362220.1.0000.5283, the protocol required written informed consent from participants, who were thoroughly briefed on the study’s objectives, procedures, and potential risks. Participation was entirely voluntary, with the option to withdraw at any point without repercussions. These measures upheld the highest ethical standards, safeguarding the rights and dignity of all involved.

### Data collection and analysis

Data collection at screening centers in São Gonçalo, Rio de Janeiro, included key sociodemographic details, comorbidities, COVID-19 symptoms, vaccination history, and test results. Symptoms such as fever, cough, loss of smell or taste, and shortness of breath were recorded alongside their onset dates. COVID-19 testing employed the RT-LAMP technique, and vaccination data encompassed the number of doses, vaccine type, and administration dates. Healthcare professionals adhered to strict safety and ethical protocols, ensuring participant consent and data confidentiality.

Statistical analyses were conducted using RStudio (version 2023.06.1 + 524) with R (version 4.2.2). Post-stratification weights were calculated to adjust for the known population distributions of ethnicity and sex in São Gonçalo (IBGE 2024), addressing potential sampling bias. Weighted generalized linear models analyzed the associations between self-reported symptoms and vaccination status of SARS-Cov-2 positive patients, as well as possible interactions with patient sex, with results reported as log odds ratio (_log_OR) and 95% confidence intervals (95% CI) to ensure robust interpretation. Statistical significance was set at a p-value < 0.05 to reduce the chance of random associations.

## Results and discussion

Unvaccinated individuals demonstrated significant positive associations with several symptoms. For fever greater than 38.5 ºC (FEVER_grt), the _log_OR was 3.64 (95% CI: 1.52 to 5.77, p = 0.0008), and for rhinitis, the _log_OR was 2.94 (95% CI: 0.82 to 5.05, p = 0.0065), indicating a higher likelihood of reporting these symptoms compared to vaccinated individuals (Figures 1b and 1c). Unvaccinated individuals also had significant positive associations with headache (_log_OR = 2.17, 95% CI: 0.06 to 4.28, p = 0.0436) and myalgia (_log_OR = 3.25, 95% CI: 1.17 to 5.34, p = 0.0023) (Figures 1g and 1i). Interestingly, for cough, unvaccinated individuals showed a significant negative association, with a _log_OR of -2.44 (95% CI: -4.53 to -0.34, p = 0.0226) (Figure 1j).

**Figure 1.**
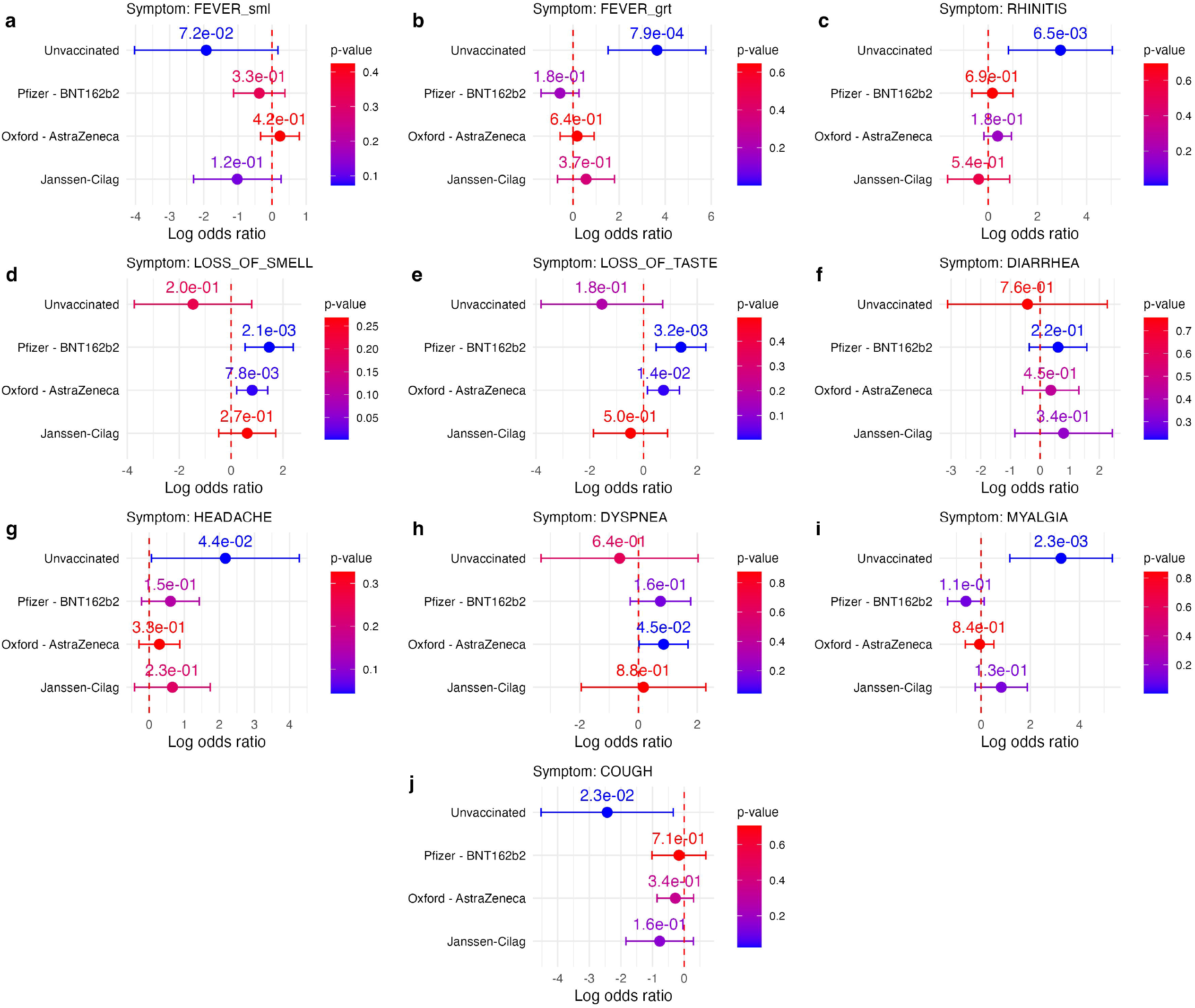
Regression estimates for associations between symptom and vaccination status. Log odds ratios (_log_OR) for the likelihood of reporting specific COVID-19 symptoms among unvaccinated individuals and those vaccinated with Pfizer-BNT162b2, Oxford-AstraZeneca, and Janssen-Cilag vaccines. Each panel corresponds to a symptom: (a) Fever (small), (b) Fever (great), (c) Rhinitis, (d) Loss of smell, (e) Loss of taste, (f) Diarrhea, (g) Headache, (h) Dyspnea, (i) Myalgia, and (j) Cough. The x-axis represents the _log_OR, where the vertical dashed red line indicates no association (_log_OR of 0). Associations are considered negative if the higher bound of the confidence interval (CI) is completely to the left of the dashed line and positive if the lower bound of the CI is completely to the right. Points and error bars represent regression estimates (_log_OR) with 95% confidence intervals, while the blue-to-red color gradient of the points reflects the p-value. Annotated p-values are shown at the top of each plot point.

Among vaccinated groups, specific associations were also observed. For loss of smell, individuals vaccinated with Oxford-AstraZeneca showed a significant positive association (_log_OR = 0.81, 95% CI: 0.21 to 1.41, p = 0.0078), as did individuals vaccinated with Pfizer-BNT162b2 (_log_OR = 1.47, 95% CI: 0.54 to 2.40, p = 0.0021) (Figure 1d). For loss of taste, a significant positive association was found in individuals vaccinated with Oxford-AstraZeneca (_log_OR = 0.75, 95% CI: 0.15 to 1.34, p = 0.0142) (Figure 1e). Additionally, for dyspnea, individuals vaccinated with Oxford-AstraZeneca demonstrated a significant positive association, with a _log_OR of 0.85 (95% CI: 0.02 to 1.69, p = 0.0449) (Figure 1h). These results (also shown in the Supplementary table I) highlight distinct symptom associations depending on vaccination status in SARS-Cov-2 positive patients.

[**Figure 1**. Regression estimates for associations between symptom and vaccination status.]

Here, interactions between vaccines and sex were also investigated, and results are summarized in the Supplementary table II. Sex differences, such as the reduced odds of dyspnea, loss of smell, and loss of taste in males with specific vaccines like Janssen might reflect a combination of behavioral, environmental, and baseline physiological differences that exacerbate susceptibility to severe immune responses. These differences seem to be especially important for the population with 50 years of age or older (Sze et al. 2020, Barek et al. 2020).

The results reveal that unvaccinated individuals demonstrate significantly higher odds of experiencing severe symptoms of SARS-CoV-2 infection, such as fever greater than 38.5 °C (_log_OR = 3.64), rhinitis (_log_OR = 2.94), headache (_log_OR = 2.17), and myalgia (_log_OR = 3.25). These associations align with prior evidence indicating that the absence of vaccination increases the risk of intense systemic and respiratory symptoms, potentially linked to heightened viral loads and unregulated inflammatory responses (Polack et al. 2020, Baden et al. 2021). Interestingly, unvaccinated individuals exhibited a significant negative association with cough (_log_OR = -2.44), which may be influenced by behavioral factors among those who chose not to receive the vaccine, such as differing health-seeking behaviors or varying levels of adherence to reporting symptoms (Chang 2021).

Among vaccinated individuals, symptom associations differed by vaccine type. Oxford-AstraZeneca and Pfizer-BNT162b2 vaccines were both associated with a higher likelihood of loss of smell (_log_OR = 0.81 and 1.47, respectively) and taste (_log_OR = 0.75 for Oxford-AstraZeneca). These sensory symptoms could reflect the vaccines’ partial attenuation of severe symptoms while not fully preventing viral effects localized to the olfactory and gustatory systems, as noted in studies examining post-vaccine symptomatology (Ayoubkhani et al. 2022). Furthermore, Oxford-AstraZeneca was uniquely associated with dyspnea (_log_OR = 0.85), highlighting the heterogenous effect of different imunization strategies on reinfection and long-COVID scenarios (Notarte et al. 2022).

These findings highlight the dual role of vaccination in mitigating severe disease while shaping the symptom profile in breakthrough cases. They emphasize the need for ongoing research into vaccine-specific immune responses and their implications for clinical management.

## Data Availability

All data produced in the present study are available upon reasonable request to the authors.

## Author contributions

RRC contributed to the conceptualization and data collection, drafted the manuscript, reviewed and edited the manuscript, and created visualizations. HRF performed data analysis and methodology development, reviewed and edited the manuscript, acquired funding, and conducted formal analysis. SVC was involved in conceptualization, drafting and editing the manuscript, supervising the project, managing project administration, acquiring funding, obtaining ethical approval, and validating the results. This collaborative effort ensured the study was thoroughly designed, effectively executed, and accurately presented.

## Ethical approval

The present research protocol was reviewed and approved by the Research Ethics Committee of the University of Grande Rio (CEP/UNIGRANRIO) under CAAE number 32362220.1.0000.5283.

## Figure legends

**Supplementary figure 1.**
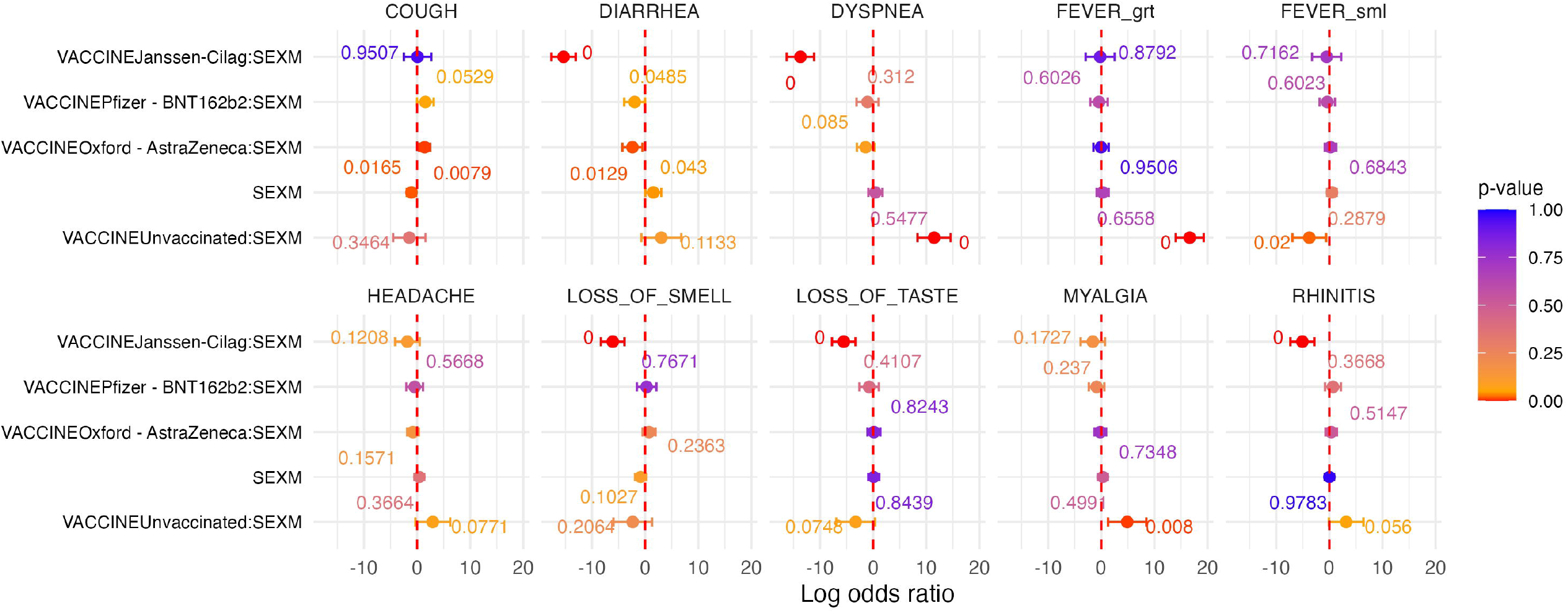
Positive weighted sex interactions in vaccine-symptom associations. Faceted forest plots illustrating the interaction effects of vaccine type and sex on reported symptoms, with log odds ratio (_log_OR) values and 95% confidence intervals displayed along the x-axis. Each facet represents a specific symptom, with points colored by p-value using a gradient, and p-values below 0.05 shown in red above the corresponding points. Significant interaction effects include notable associations for unvaccinated males with symptoms such as fever > 38.5□°C (_log_OR: 16.65, CI: [14.02, 19.28]), dyspnea (_log_OR: 11.46, CI: [8.34, 14.59]), and myalgia (_log_OR: 4.88, CI: [1.28, 8.48]). Negative associations are observed for Janssen-Cilag vaccine interactions with symptoms like dyspnea (_log_OR: -13.70, CI: [-16.28, -11.12]), diarrhea (_log_OR: -15.37, CI: [-17.66, -13.07]), and loss of smell (_log_OR: -6.10, CI: [-8.32, -3.89]), indicating reduced odds of these symptoms.

