## Supplementary table I for "Weighted analysis of symptom profile by vaccination status in positive SARS-CoV-2 cases: observational study in São Gonçalo, Brazil"

Tables

**Supplementary table I**. Weighted regression analysis of vaccine-symptom associations.

| Symptom | Variable | logOR | CI_Lower | CI_Upper | p-value |
|---|---|---|---|---|---|
| FEVER_grt | Unvaccinated | 3.64 | 1.52 | 5.77 | 0.0008*** |
| MYALGIA | Unvaccinated | 3.25 | 1.17 | 5.34 | 0.0023** |
| RHINITIS | Unvaccinated | 2.94 | 0.82 | 5.05 | 0.0065** |
| HEADACHE | Unvaccinated | 2.17 | 0.06 | 4.28 | 0.0436* |
| LOSS_OF_SMELL | Pfizer - BNT162b2 | 1.47 | 0.54 | 2.40 | 0.0021** |
| LOSS_OF_TASTE | Pfizer - BNT162b2 | 1.40 | 0.47 | 2.32 | 0.0032** |
| DYSPNEA | Oxford - AstraZeneca | 0.85 | 0.02 | 1.69 | 0.0449* |
| MYALGIA | Janssen-Cilag | 0.82 | -0.24 | 1.88 | 0.1278 |
| LOSS_OF_SMELL | Oxford - AstraZeneca | 0.81 | 0.21 | 1.41 | 0.0078** |
| DIARRHEA | Janssen-Cilag | 0.79 | -0.85 | 2.44 | 0.3443 |
| LOSS_OF_TASTE | Oxford - AstraZeneca | 0.75 | 0.15 | 1.34 | 0.0142* |
| DYSPNEA | Pfizer - BNT162b2 | 0.75 | -0.28 | 1.78 | 0.1561 |
| HEADACHE | Janssen-Cilag | 0.66 | -0.42 | 1.74 | 0.2294 |
| LOSS_OF_SMELL | Janssen-Cilag | 0.62 | -0.48 | 1.72 | 0.2682 |
| DIARRHEA | Pfizer - BNT162b2 | 0.61 | -0.37 | 1.58 | 0.2220 |
| HEADACHE | Pfizer - BNT162b2 | 0.61 | -0.22 | 1.43 | 0.1491 |
| FEVER_grt | Janssen-Cilag | 0.56 | -0.67 | 1.80 | 0.3727 |
| RHINITIS | Oxford - AstraZeneca | 0.38 | -0.18 | 0.94 | 0.1808 |
| DIARRHEA | Oxford - AstraZeneca | 0.36 | -0.59 | 1.31 | 0.4539 |
| HEADACHE | Oxford - AstraZeneca | 0.29 | -0.29 | 0.87 | 0.3271 |
| FEVER_sml | Oxford - AstraZeneca | 0.23 | -0.34 | 0.80 | 0.4240 |
| FEVER_grt | Oxford - AstraZeneca | 0.17 | -0.57 | 0.91 | 0.6450 |
| DYSPNEA | Janssen-Cilag | 0.17 | -1.95 | 2.29 | 0.8751 |
| RHINITIS | Pfizer - BNT162b2 | 0.17 | -0.67 | 1.00 | 0.6940 |
| MYALGIA | Oxford - AstraZeneca | -0.06 | -0.64 | 0.52 | 0.8429 |
| COUGH | Pfizer - BNT162b2 | -0.16 | -1.02 | 0.69 | 0.7057 |
| COUGH | Oxford - AstraZeneca | -0.28 | -0.85 | 0.30 | 0.3447 |
| FEVER_sml | Pfizer - BNT162b2 | -0.37 | -1.13 | 0.38 | 0.3306 |
| RHINITIS | Janssen-Cilag | -0.39 | -1.65 | 0.87 | 0.5449 |
| DIARRHEA | Unvaccinated | -0.42 | -3.12 | 2.27 | 0.7575 |
| LOSS_OF_TASTE | Janssen-Cilag | -0.48 | -1.86 | 0.90 | 0.4951 |
| FEVER_grt | Pfizer - BNT162b2 | -0.57 | -1.39 | 0.25 | 0.1764 |
| MYALGIA | Pfizer - BNT162b2 | -0.61 | -1.36 | 0.13 | 0.1054 |
| DYSPNEA | Unvaccinated | -0.64 | -3.32 | 2.03 | 0.6371 |
| COUGH | Janssen-Cilag | -0.77 | -1.84 | 0.29 | 0.1557 |
| FEVER_sml | Janssen-Cilag | -1.02 | -2.30 | 0.26 | 0.1197 |
| LOSS_OF_SMELL | Unvaccinated | -1.47 | -3.73 | 0.80 | 0.2039 |
| LOSS_OF_TASTE | Unvaccinated | -1.55 | -3.81 | 0.71 | 0.1801 |
| FEVER_sml | Unvaccinated | -1.93 | -4.03 | 0.17 | 0.0725 |
| COUGH | Unvaccinated | -2.44 | -4.53 | -0.34 | 0.0226* |

* = p ≤ 0.05, ** = p ≤ 0.01, *** = p ≤ 0.001.
