## Supplementary table II for "Weighted analysis of symptom profile by vaccination status in positive SARS-CoV-2 cases: observational study in São Gonçalo, Brazil"

Tables

**Supplementary table II**. Weighted interaction analysis of sex in vaccine-symptom associations.

| Symptom | Variable | logOR | CI_Lower | CI_Upper | p-value |
|---|---|---|---|---|---|
| FEVER_grt | VACCINEUnvaccinated:SEXM | 16.65 | 14.02 | 19.28 | 0.0000*** |
| DYSPNEA | VACCINEUnvaccinated:SEXM | 11.46 | 8.34 | 14.59 | 0.0000*** |
| MYALGIA | VACCINEUnvaccinated:SEXM | 4.88 | 1.28 | 8.48 | 0.0080** |
| RHINITIS | VACCINEUnvaccinated:SEXM | 3.17 | -0.08 | 6.42 | 0.0560 |
| DIARRHEA | VACCINEUnvaccinated:SEXM | 3.04 | -0.72 | 6.80 | 0.1133 |
| HEADACHE | VACCINEUnvaccinated:SEXM | 2.95 | -0.32 | 6.22 | 0.0771 |
| COUGH | VACCINEPfizer - BNT162b2:SEXM | 1.55 | -0.02 | 3.12 | 0.0529 |
| DIARRHEA | SEXM | 1.55 | 0.05 | 3.04 | 0.0430* |
| COUGH | VACCINEOxford - AstraZeneca:SEXM | 1.42 | 0.37 | 2.46 | 0.0079** |
| LOSS_OF_SMELL | VACCINEOxford - AstraZeneca:SEXM | 0.73 | -0.48 | 1.95 | 0.2363 |
| RHINITIS | VACCINEPfizer - BNT162b2:SEXM | 0.69 | -0.80 | 2.18 | 0.3668 |
| FEVER_sml | SEXM | 0.48 | -0.41 | 1.38 | 0.2879 |
| HEADACHE | SEXM | 0.42 | -0.49 | 1.34 | 0.3664 |
| DYSPNEA | SEXM | 0.41 | -0.92 | 1.73 | 0.5477 |
| RHINITIS | VACCINEOxford - AstraZeneca:SEXM | 0.35 | -0.71 | 1.42 | 0.5147 |
| MYALGIA | SEXM | 0.31 | -0.60 | 1.23 | 0.4991 |
| LOSS_OF_SMELL | VACCINEPfizer - BNT162b2:SEXM | 0.28 | -1.56 | 2.12 | 0.7671 |
| FEVER_grt | SEXM | 0.26 | -0.87 | 1.39 | 0.6558 |
| FEVER_sml | VACCINEOxford - AstraZeneca:SEXM | 0.22 | -0.85 | 1.30 | 0.6843 |
| LOSS_OF_TASTE | VACCINEOxford - AstraZeneca:SEXM | 0.14 | -1.07 | 1.35 | 0.8243 |
| LOSS_OF_TASTE | SEXM | 0.10 | -0.93 | 1.14 | 0.8439 |
| COUGH | VACCINEJanssen-Cilag:SEXM | 0.08 | -2.49 | 2.65 | 0.9507 |
| RHINITIS | SEXM | 0.01 | -0.85 | 0.88 | 0.9783 |
| FEVER_grt | VACCINEOxford - AstraZeneca:SEXM | -0.05 | -1.48 | 1.39 | 0.9506 |
| MYALGIA | VACCINEOxford - AstraZeneca:SEXM | -0.19 | -1.29 | 0.91 | 0.7348 |
| FEVER_grt | VACCINEJanssen-Cilag:SEXM | -0.21 | -2.94 | 2.52 | 0.8792 |
| FEVER_sml | VACCINEPfizer - BNT162b2:SEXM | -0.38 | -1.83 | 1.06 | 0.6023 |
| FEVER_grt | VACCINEPfizer - BNT162b2:SEXM | -0.44 | -2.09 | 1.21 | 0.6026 |
| HEADACHE | VACCINEPfizer - BNT162b2:SEXM | -0.46 | -2.03 | 1.11 | 0.5668 |
| FEVER_sml | VACCINEJanssen-Cilag:SEXM | -0.51 | -3.25 | 2.23 | 0.7162 |
| LOSS_OF_TASTE | VACCINEPfizer - BNT162b2:SEXM | -0.77 | -2.60 | 1.06 | 0.4107 |
| HEADACHE | VACCINEOxford - AstraZeneca:SEXM | -0.78 | -1.87 | 0.30 | 0.1571 |
| LOSS_OF_SMELL | SEXM | -0.86 | -1.90 | 0.17 | 0.1027 |
| MYALGIA | VACCINEPfizer - BNT162b2:SEXM | -0.88 | -2.34 | 0.58 | 0.2370 |
| COUGH | SEXM | -1.07 | -1.94 | -0.20 | 0.0165* |
| DYSPNEA | VACCINEPfizer - BNT162b2:SEXM | -1.07 | -3.14 | 1.00 | 0.3120 |
| DYSPNEA | VACCINEOxford - AstraZeneca:SEXM | -1.44 | -3.07 | 0.20 | 0.0850 |
| COUGH | VACCINEUnvaccinated:SEXM | -1.47 | -4.52 | 1.59 | 0.3464 |
| MYALGIA | VACCINEJanssen-Cilag:SEXM | -1.59 | -3.88 | 0.70 | 0.1727 |
| HEADACHE | VACCINEJanssen-Cilag:SEXM | -1.83 | -4.15 | 0.48 | 0.1208 |
| DIARRHEA | VACCINEPfizer - BNT162b2:SEXM | -1.98 | -3.94 | -0.01 | 0.0485* |
| LOSS_OF_SMELL | VACCINEUnvaccinated:SEXM | -2.34 | -5.97 | 1.29 | 0.2064 |
| DIARRHEA | VACCINEOxford - AstraZeneca:SEXM | -2.39 | -4.28 | -0.51 | 0.0129* |
| LOSS_OF_TASTE | VACCINEUnvaccinated:SEXM | -3.30 | -6.93 | 0.33 | 0.0748 |
| FEVER_sml | VACCINEUnvaccinated:SEXM | -3.77 | -6.94 | -0.60 | 0.0200* |
| RHINITIS | VACCINEJanssen-Cilag:SEXM | -5.07 | -7.33 | -2.82 | 0.0000*** |
| LOSS_OF_TASTE | VACCINEJanssen-Cilag:SEXM | -5.54 | -7.76 | -3.33 | 0.0000*** |
| LOSS_OF_SMELL | VACCINEJanssen-Cilag:SEXM | -6.10 | -8.32 | -3.89 | 0.0000*** |
| DYSPNEA | VACCINEJanssen-Cilag:SEXM | -13.70 | -16.28 | -11.12 | 0.0000*** |
| DIARRHEA | VACCINEJanssen-Cilag:SEXM | -15.37 | -17.66 | -13.07 | 0.0000*** |

* = $p \leq 0.05$, ** = $p \leq 0.01$, *** = $p \leq 0.001$.
